# A Model Describing COVID-19 Community Transmission Taking into Account Asymptomatic Carriers and Risk Mitigation

**DOI:** 10.1101/2020.03.18.20037994

**Authors:** Jacob B. Aguilar, Jeremy Samuel Faust, Lauren M. Westafer, Juan B. Gutierrez

## Abstract

Coronavirus disease 2019 (COVID-19) is a novel human respiratory disease caused by the SARS-CoV-2 virus. Asymptomatic carriers of the COVID-19 virus display no clinical symptoms but are known to be contagious. Recent evidence reveals that this subpopulation, as well as persons with mild disease, are a major contributor in the propagation of the disease. The rapid spread of COVID-19 forced governments around the world to establish and enforce generalized risk mitigation strategies, from lockdowns to guidelines for social distancing, in an effort to minimize community transmission. This created an unprecedented epidemiological situation not properly characterized by existing mathematical models of isolation and quarantine. In this manuscript, we present a mathematical model for community transmission of COVID-19 taking into account asymptomatic carriers and varying degrees of risk mitigation. The main results consist of an exact calculation of the effective reproduction number 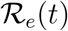, and a modeling framework that enables the quantification of the effect of risk mitigation and asymptomatism on community transmission. A computation of 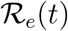 is provided using mean parameters. The point estimate of the basic reproduction number is 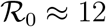.

## 1 Background

Coronavirus disease 2019 (COVID-19) is a novel human respiratory disease caused by the SARS-CoV-2 virus. The first cases of COVID-19 disease surfaced during late December 2019 in Wuhan city, the capital of Hubei province in China. Shortly after, the virus quickly spread to several countries *(1)*. On January 30, 2020 The World Health Organization (WHO) declared the virus as a public health emergency of international scope *(2)*. Forty one days later, on March 11, 2020 it was officially declared to be a global pandemic *(3)*.

Asymptomatic individuals in the context of COVID-19 disease are subjects who carry a viral load, but do not show clinical symptoms. When the first cases appeared in China, there was no clarity about the existence of asymptomatic carriers. The evolution of our understanding of this matter has produced a very broad span of estimates, ranging from 1% to 88%, as summarized next:

- 1.1% from the Chinese Center for Disease Control, cross sectional study *(4)*. The criteria for inclusion was presence of symptoms. The only surprise is that this study found any asymptomatic carrier at all.
- 17.9% from the Princess cruise ship in Japan *(5)*. However, the age pyramid of passengers was heavily biased toward > 60 year-old, precisely the age group most likely to develop symptoms. This finding does not translate to communities.
- 30.8% from Japanese evacuees from Wuhan *(6)*. The age pyramid of this group was also tilted toward seniority.
- 50-75% from Vo’Euganeo in Italy. An entire village was tested *(7)*.
- +80% from reanalysis of Chinese data *(8)*.
- 88% in pregnant women in a maternity ward were positive for SARS-CoV-2 upon admission but had no symptoms of COVID-19 at presentation *(9)*.

Asymptomatic carriers pose a silent threat to communities because these individuals might not adhere to risk mitigation strategies (e.g. wearing face masks). Asymptomatic and symptomatic carriers may have similar levels of viral load and infectiousness *(10,11)*. Since they are frequently undetected by public health systems, the potential for sustained contagion is high *(5,12)*.

The rapid spread of COVID-19 forced governments around the world to establish and enforce generalized risk mitigation strategies, from lockdowns to guidelines for social distancing, in an effort to minimize community transmission *(13-15)*. From a mathematical point of view, the effects resulting from varying degrees of risk mitigation are not captured by existing quarantine models whose formulation depends on the isolation of a given sub-population, whereas in COVID-19 entire societies were subject to restrictions *(16-19)*.

The primary aim of this manuscript is to qualitatively characterize the epidemiological dynamics of SARS-CoV-2 via a compartmentalized model that takes into account the asymptomatic sub-population and risk mitigation conditions. This manuscript is organized as follows: Sect. 2.1 presents a generalized model in which the additional models covered in this manuscript are specific cases and modifications, Sect. 2.2 features a slight simplification of the model featured in Sect. 2.1 with a reproduction number which admits a natural biological interpretation and a numerical implementation, Sect. 3 builds upon the simplified model presented in Sect. 2.2 through means of a modification which takes into account the changes in behavioral patterns during the risk mitigation period which resulted in a reduction of the susceptible population and contains a numerical plot of the effective reproduction number of the model, Sect. 4 is focused on covering the biological relevance of the reproduction number listed in Sect. 2.2, Sect. 5 is the conclusion and Sect. 6 contains tables in which the biological and computationally determined parameters are listed separately.

## 2 Mathematical Models

This section contains variations of models which fall into the class of models covered by Aguilar and Gutierrez (2020) (20). The *SEYAR* model for the spread of COVID- 19 is formulated by decomposing the total host population (*N*) into the following five epidemiological classes: susceptible human (*S*), exposed human (*E*), symptomatic human (*Y*), asymptomatic human (*A*), and recovered human (*R*).The following generalized SEYAR dynamical system, is given by Equation 1 below, (see Figure 1 below):

**Figure 1:**
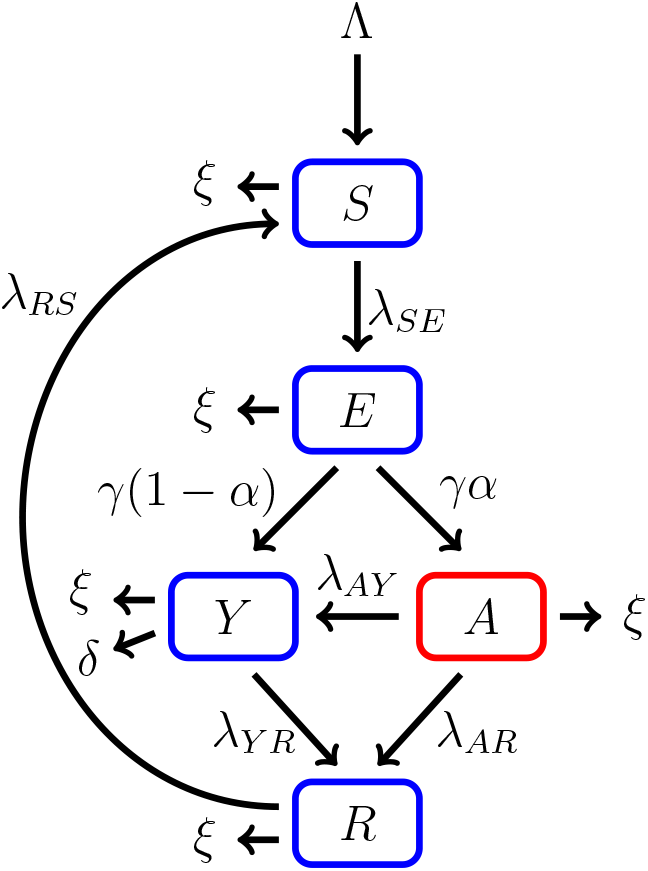
This figure is a schematic diagram of a generalized COVID-19 model including an asymptomatic compartment. The longer arrows represent progression from one compartment to the next. Hosts enter the susceptible compartment either through birth of migration and then progress through each additional compartment subject to the rates described above.

### 2.1 Generalized Model

The following generalized SEYAR dynamical system, is given by Equation 1 below, (see Figure 1 below):

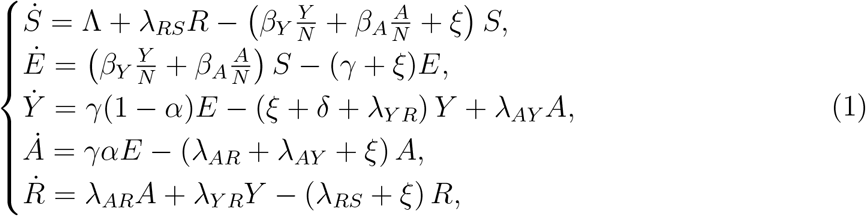

where *N* = *S* + *E* + *Y* + *A* + *R*. The demographic parameters Λ and ξ denote the human recruitment and mortality rates, respectively. While λ*_AY_* and λ*_RS_* are the asymptomatic to symptomatic transition and relapse rates, respectively. It is worth mentioning that for a basic SEIR model, where there is only one infected compartment, the progression rate from the susceptible to the exposed class λ*_SE_* is equal to the product of the effective contact rate *β* and the proportion of infected individuals 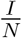. The force of infection is

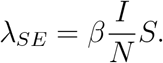

In our model, we decompose the infected compartment into symptomatic and asymptomatic sub-compartments. Due to this decomposition, the force of infection is given by the weighted sum

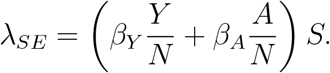

Disease-Free Equilibrium (DFE) points are solutions of a dynamical system corresponding to the case where no disease is present in the population. The reproduction number 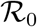 is a threshold value that characterizes the local asymptotic stability of the underlying dynamical system at a disease-free equilibrium. Listed below in Lemma 1 is a proof of the reproduction number associated to the generalized Model 1.

#### Lemma 1.

*(Reproduction Number for the SEYAR COVID-19 Model). Define the following quantity*

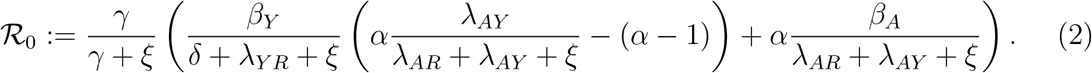

*Then, the DFE* w^⋆^ *for the SEYAR model in Equation 1 is locally asymptotically stable provided that* 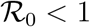 *and unstable if* 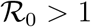.

#### Proof.

We order the compartments so that the first four correspond to the infected sub-populations and denote **w** = (*E*, *Y*, *A*, *R*, *S*)*^T^*. The corresponding DFE is

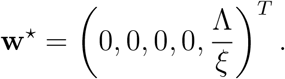

The system in Equation 1 can be rewritten using the next generation method *(21)* as 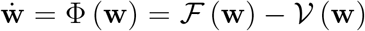, where 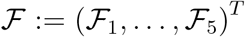 and 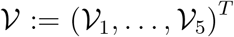, or more explicitly

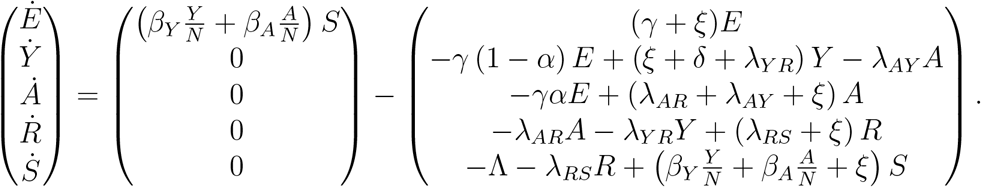

The matrix 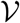 admits the decomposition 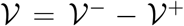, where the component-wise definition is inherited. In a biological context, 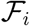 is the rate of appearance of new infections in compartment *i*, 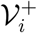 stands for the rate of transfer of individuals into compartment *i* by any other means and 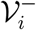 is the rate of transfer of individuals out of compartment *i*. Now, let *F* and *V* be the following sub-matrices of the Jacobian of the above system, evaluated at the solution **w**^⋆^

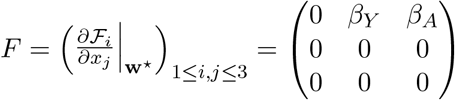

and

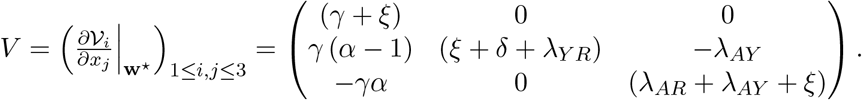

A direct calculation shows that

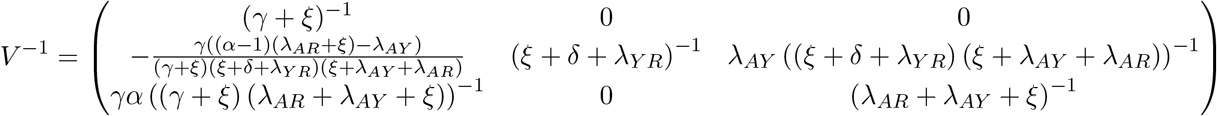

and *FV*^−1^ is given by the following matrix

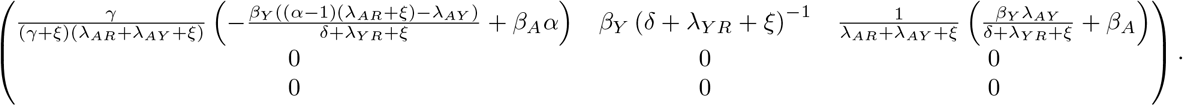

Let 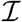 denote the 3 × 3 identity matrix, so that the characteristic polynomial *P*(λ) of the matrix *FV*^−1^ is given by

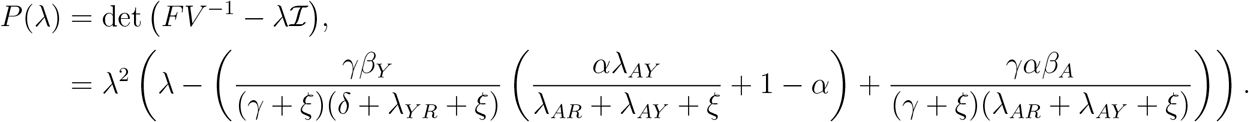

The solution set {*λ_i_*}_1≤_*_i_*_≤3_ is given by

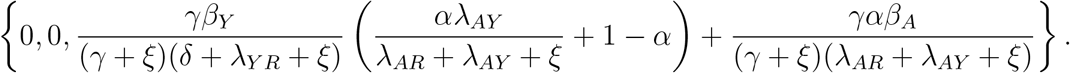

Therefore, the reproduction number for the *SEYAR* model in Equation 1 is given by

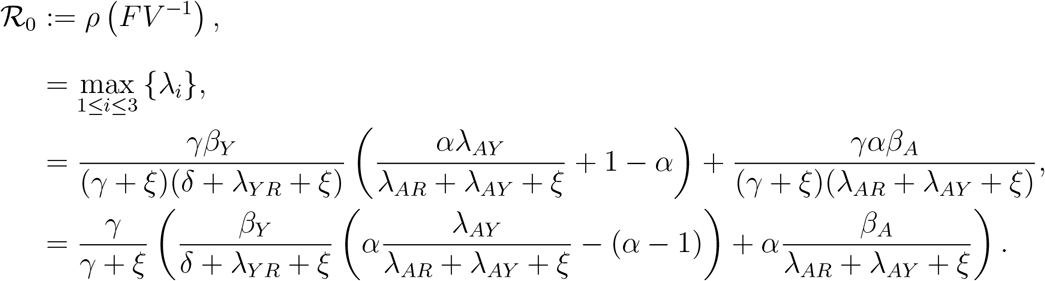

The proof of the lemma regarding the local asymptotic stability of the DFE **w**^⋆^ corresponding to the *SEYAR* model in Equation 1 is now complete since **w**^⋆^ is locally asymptotically stable if 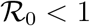, but unstable if 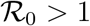 *(21, Theorem 2*).

### 2.2 Simplified Model

This section features a simplification of Model 1 corresponding to the absence of demographic parameters, the asymptomatic to symptomatic transition rate and the relapse rate.

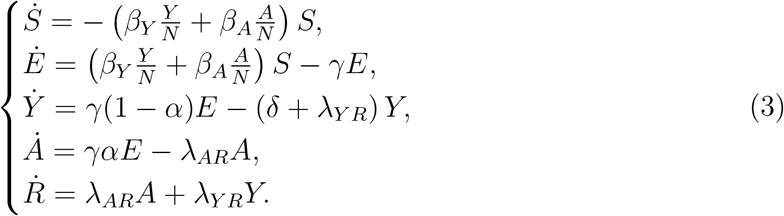

**Figure 2:**
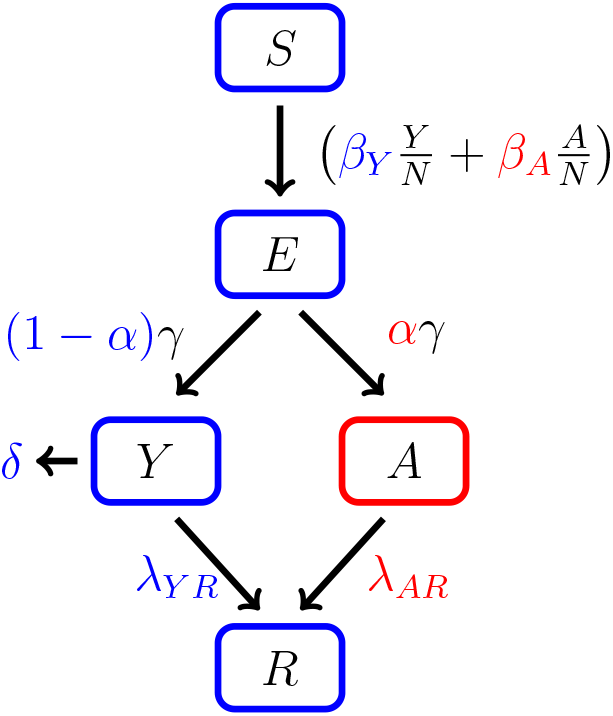
Schematic diagram of a COVID-19 model including an asymptomatic compartment. The arrows, except the disease-induced death (*δ*), represent progression from one compartment to the next. Hosts progress through each compartment subject to the rates described below.

The reproduction number arising from the dynamical system 3 is given by the following equation

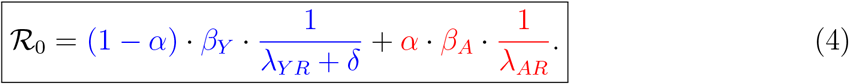

The reproduction number featured in Equation 4 above corresponds to a DFE solution given by **v**^⋆^ = (0, 0, 0, 0, *S*_0_)*^T^* and the absence of demographic parameters and the asymptomatic to symptomatic transition rate. It can be alternatively obtained by letting

**Table 1:** Average values of parameters used to compute Figures 4 and 5. See Appendix for sources. The fitted parameters were selected from the optimal fit of a calibrated model for the City of San Antonio, TX (data not shown).

| Parameter | Description | Dimension | Value |  |
| --- | --- | --- | --- | --- |
| *FITTED* |  |  |  |  |
| $\beta_Y$ | Effective contact rate from symptomatic to susceptible. | $days^{-1}$ | 1.17 | |
| $\beta_A$ | Effective contact rate from asymptomatic to susceptible. | $days^{-1}$ | 1.16 | |
| $k$ | Risk mitigation coefficient | n/a | 0.03 | |
| Parameter | Description | Dimension | Mean | Variance |
| *BIOLOGICAL* |  |  |  |  |
| $\gamma^{-1}$ | Mean latent period. | days | 5.2 | 0.5 |
| $\alpha$ | Probability of becoming asymptomatic upon infection. | n/a | 0.60 | 0.10 |
| $\lambda_{YR}^{-1}$ | Mean symptomatic infectious period. | $days$ | 13.5 | 31.8 |
| $\lambda_{AR}^{-1}$ | Mean asymptomatic infectious period. | $days$ | 8.33 | n/a |
| $\delta$ | Disease-induced death rate. | $days^{-1}$ | 0.026 | 0.094 |

The calculation of 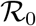 during the first stages of an epidemic poses significant challenges. Evidence of this difficulty was observed in the 2009 influenza A (H1N1) virus pandemic *(22)*. Particularly, the COVID-19 pandemic has a different characterization in each country in which it has spread due to differences in surveillance capabilities of public health systems, socioeconomic factors, and environmental conditions.

The first three weeks of community transmission is well characterized by an exponential function in multiple locations. Figure 3 shows the number of cases reported in thirteen countries with universal health care and strong surveillance systems until March 25, 2020. Ten of these countries are in the European zone, plus Australia, Canada and Japan. An exponential fitting for each country reveals an average coefficient of determination *R*^2^ = 0.9846 ± 0.0164. The average growth rate r in the exponential model *Y* = *a* · (1 + *r*)*^t^*, where *t* is time measured in days, is *r* = 23.32%, and the average of the initial conditions is *a* = 103 cases. Thus,

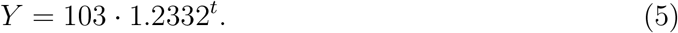

**Figure 3:**
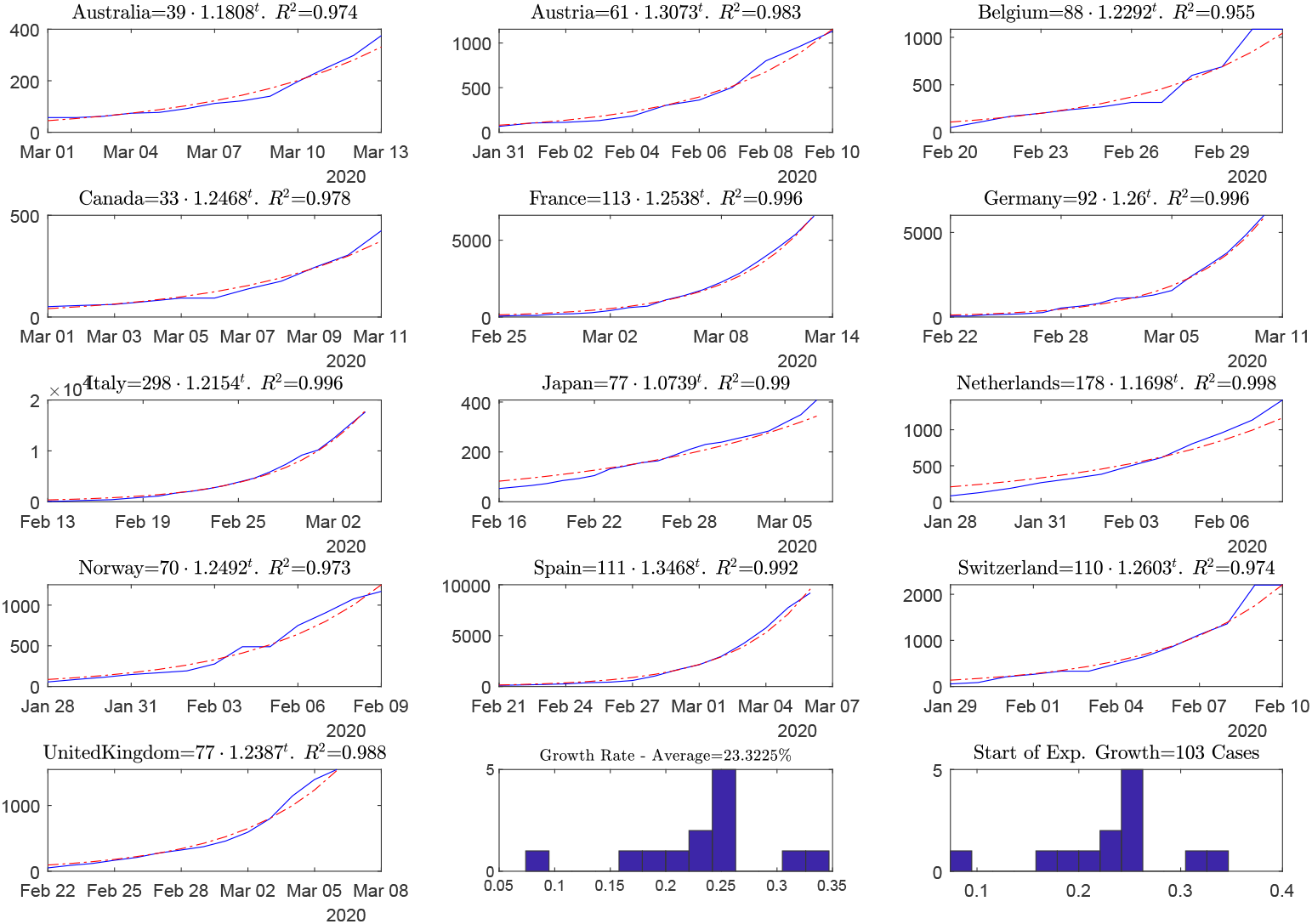
First three weeks (or less) of data for thirteen countries with COVID-19 cases and strong surveillance systems for communicable diseases.

There are well known challenges in attempting to fit an exponential function to epidemiological data *(23-25)*. To compare the output of the model to the data from the thirteen countries studied, the growth rate found in Equation 5 was superimposed on the model. The initial condition *a*_0_ in the exponential function *Y* = *a*_0_ · (1 + *r*)*^t^* was fitted to the dynamical system with the Nelder-Meade simplex algorithm (26).

Figure 4 shows a calculation of System 3 using the parameter values listed in Table 1. This representation of the dynamics of the disease must be understood as a theoretical development; in reality, the progression of an epidemic depends on a multitude of factors that necessarily result in deviations from this ideal case.

**Figure 4:**
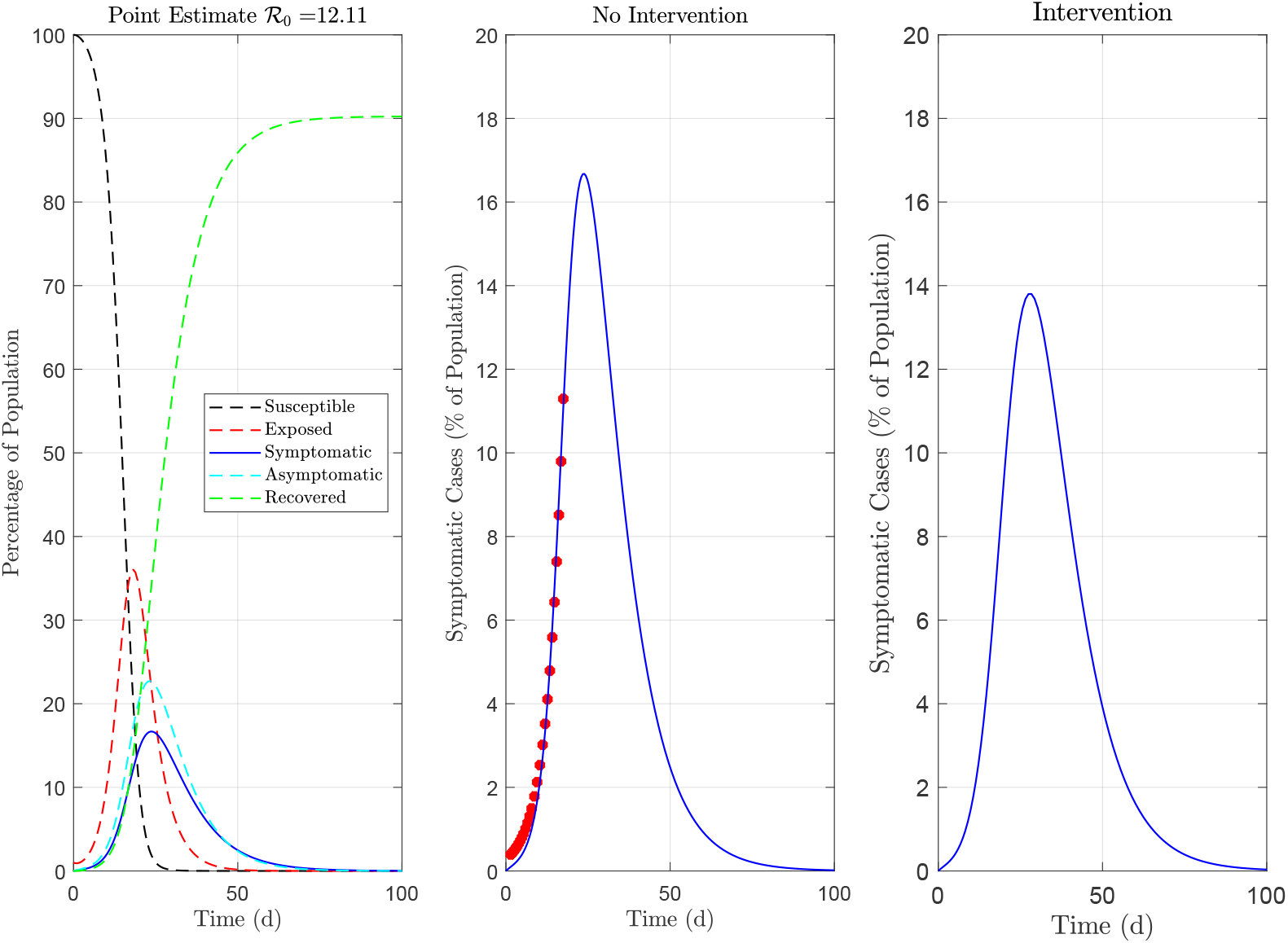
Numerical implementation of System 3 with parameter values listed in Table 1. The left-most panel shows the time series corresponding to a point estimate of 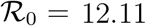. The center panel shows a times series of the symptomatic compartment; the red dots represent the exponential function whose parameters are the average of the thirteen countries studied. The right-most panel shows a simulation representing the effect of limiting contact between the susceptible and infected populations. At the time of writing there is no data available to calibrate an intervention model.

## 3 Modeling Risk Mitigation Conditions

Changes in behavioral patterns in response to an outbreak have an effect on the spread of a given disease. When an infectious pathogen threatens a community, individual awareness and public health interventions can motivate a portion of community individuals to take measures to reduce their exposure to the pathogen. The rapid spread and high contagiousness of COVID-19 resulted in exponential growth during the first three weeks of the outbreak as shown in Figure 3 of Section 2.2. In response, drastic measures were taken by community leaders in order to reduce the susceptiblity of the population and, as a result, slow down the spread of the disease. Social distancing, the cancellation of events likely to attract crowds, the closing of schools and working from home will all have a drastic impact on the size of the susceptible population at a given time *(27, 28)*.

Let *Q*(*t*) denote the risk at time *t*. *Q*(*t*) should be chosen such that the rate of change of risk decreases proportionally to the amount of risk present, i.e. the more danger there is, the more careful people are. Consider the following initial value problem:

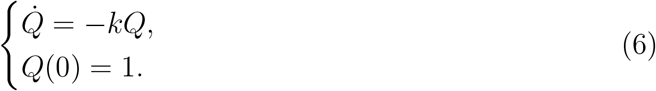

The solution is given by *Q*(*t*) = *e*^−^*^kt^*. This framework captures a broad spectrum of risk mitigation strategies, from shelter-in-place orders (e.g. *k* = 0.05), to minimal guidelines for social distancing (e.g. *k* ≈ 0). Thus, the following model modification is proposed to model the effect of these behavioral changes

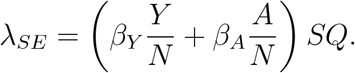

In the above, 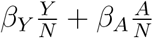 *S* describes the infection force of the disease and *Q*(*t*) = *e*−*^kt^* measures the risk due to the behavioral change of the susceptible individuals. Therefore, we arrive at the following alteration of System 3.

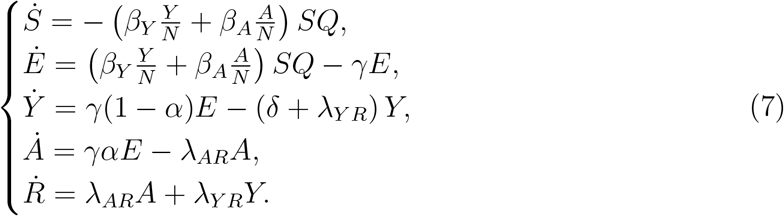

Next, we provide a formula for the effective reproduction number of System 7. In particular, we have Theorem 1 below.

### Theorem 1.

*The effective reproduction number corresponding to the dynamical system given by Equation 7 is given by the following equation*

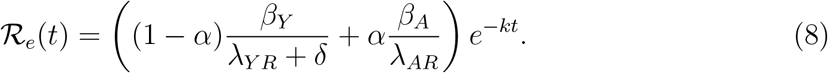

### Proof.

The matrix *V* corresponding to System 7 is unchanged by the modification involving the exponential multiplier *Q*(*t*):= *e*^−^*^kt^*. Let *B_Y_*(*t*):= *β_Y_Q*(*t*) and *B_A_*(*t*):= *β_A_Q*(*t*), then

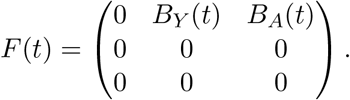

In the constant case, i.e. *B_Y_*(*t*) ≡ *β_Y_* and *B_A_*(*t*) ≡ *β_A_* for all *t* ≥ 0, we have

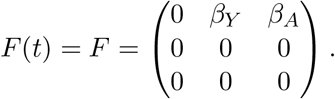

Thus, it follows that

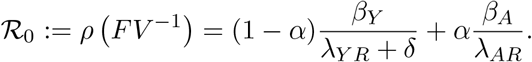

It is clear that the exponential multiplier *Q* preserves the non-negativity of *F*(*t*) for all *t*. For fixed time *t* = *t*_0_, the quantity *ρ* (*F*(*t*_0_)*V*^−1^) is equal to the maximum eigenvalue of the linear operator *F*(*t*_0_)*V*^−1^.

For *t* ∈ ℝ_+_, the eigenvalue functions depend continuously on the coefficients of the characteristic polynomial. The high regularity of the exponential multiplier measuring the risk effect, i.e. 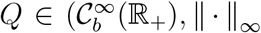 (the space of smooth bounded functions equipped with the supremum norm) in combination with its monotonic decreasing behavior for all *t* ≥ 0 ensures that the ordering of the eigenvalue functions is preserved and the maximum is well-defined. Precisely, denote dim(*F*(*t*)*V*^−1^) = *n*, the spectral radius function *ρ* (*F*(*t*)*V*^−1^) is interpreted to be the largest eigenvalue function λ*_i_*(*t*) for *i* = 1, ⋯, *n* in the Banach space *L*^∞^(ℝ_+_).

In this case, the spectral radius function *ρ* (*F*(*t*)*V*^−1^) is well-defined and is given by:

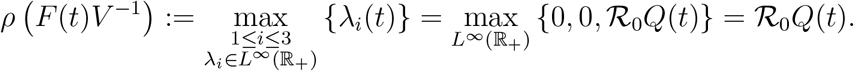

Therefore, we arrive at the following reproduction function which takes into account an exponential decline in the susceptible population:

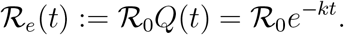

### Remark 3.1.

*It is a direct consequence of Equation 8 that* 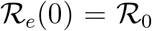 *and the disease should stop spreading after* 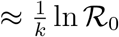 *days provided continual isolation resulting in such a drastic decrease in susceptibility is maintained. Particularly, it follows that* 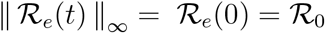 *and* 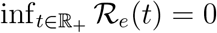, *thus* 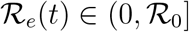 *for all t* ∈ ℝ_+_.

*In general, the susceptible population decreases in response to the infection force which is reflected in the canonical definition of* 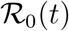, *as given by equation 10. The above modification accounts for a decline in the susceptible population that is consistent with observable results pertaining to the COVID-19 pandemic*.

The modified *SEYAR* model 7 can be used as a tool to explore multiple scenarios corresponding to different interventions. Featured below is a plot of the effective reproduction number 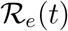 labeled as Equation 8.

**Figure 5:**
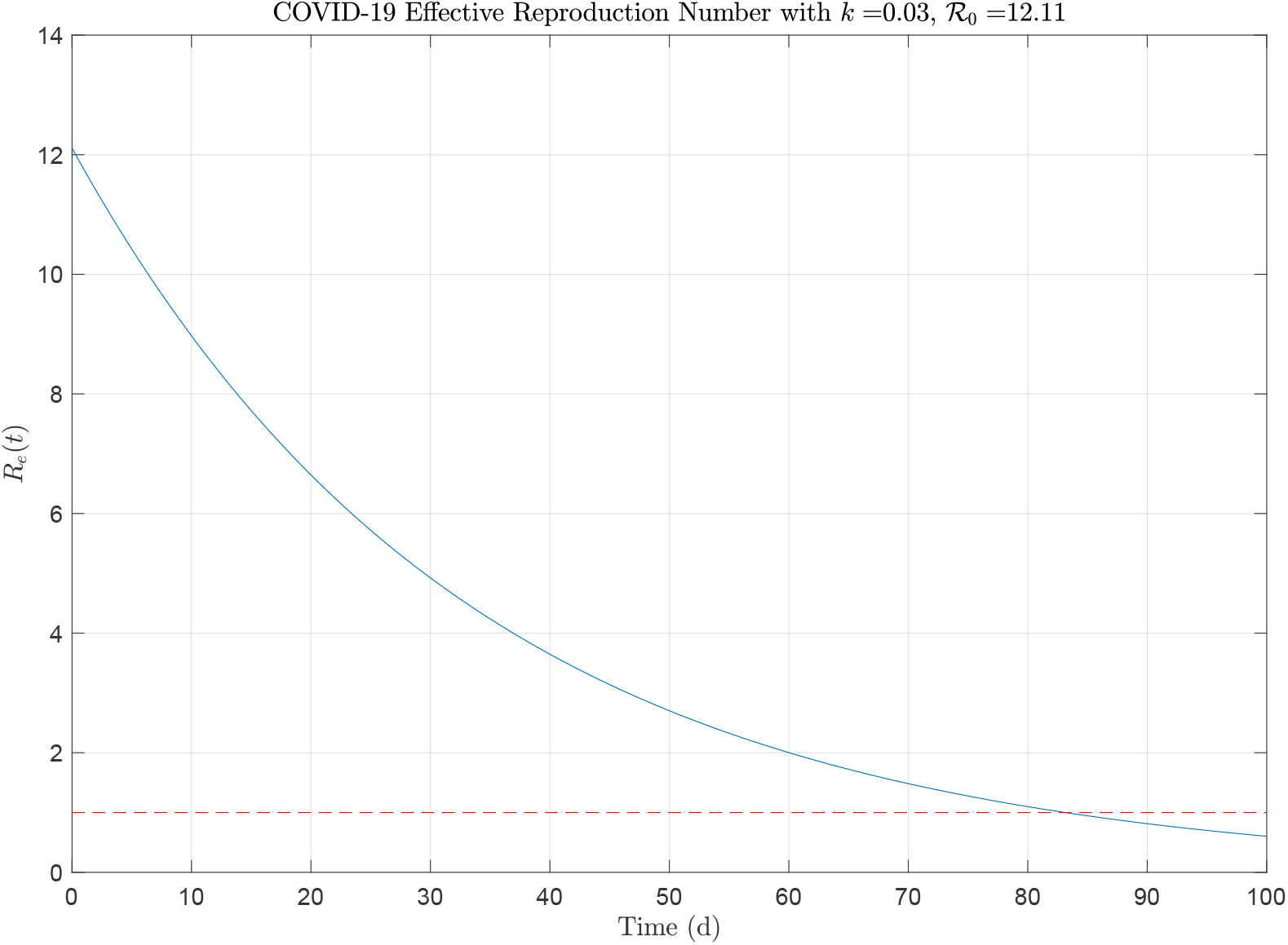
The effective reproduction number given by Equation 8 was computed with parameter values listed in Table 1. The red dashed line corresponds to 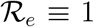. Notice that the function 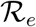 decreases below 1 after 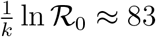 days, as mentioned in Remark 3.1.

## 4 Discussion

The reproduction number 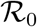 shown in Equation 4 arising from the simplified model *(3)* admits a natural biological interpretation. To guide this discussion, it is pertinent to refer to the original epidemic model proposed by W. O. Kermack and A. G. McKendrick in 1927 *(29)*, see Figure 6 below. The corresponding dynamical system is given by

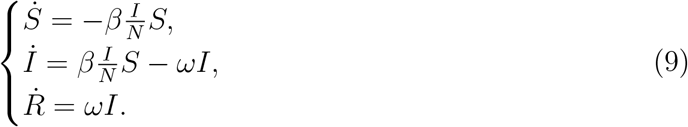

**Figure 6:**
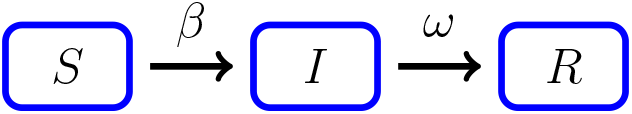
This figure is a schematic diagram of a *SIR* model consisted of three compartments, namely: susceptible (*S*), infected (*I*) and recovered (*R*). Humans progress through each compartment subject to the rates described above.

Epidemiologically speaking, the basic reproduction number is the average number of secondary infections generated by a single infection in a completely susceptible population. It is proportional to the product of infection/contact (*a*), contact/time (*b*) and time/infection (*c*). The quantity a is the infection probability between susceptible and infectious individuals, *b* is the mean contact rate between susceptible and infectious individuals and *c* is the mean duration of the infectious period.

The case of an increasing infected sub-population corresponds to the occurrence of an epidemic. This happens provided that 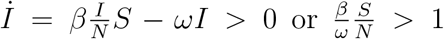. Under the assumption that in the beginning of an epidemic, virtually the total population is susceptible, that is 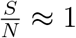. As a result, we arrive at the following equivalent condition

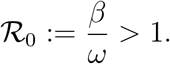

The parameter *β* in Figure 6 is equal to *ab* and *ω* is equal to *c*^−1^. This combination of parameters stands to reason as it is a ratio of the effective contact rate *β* and the mean infectious period *ω*^−1^.

Since the case fatality ratio is of negligible size (i.e. *δ* ≈ 0), the reproduction number featured in Equation 4 has a similar natural biological interpretation as the sum of ratios consisting of the effective contact rates *β_Y_, β_A_* and mean infectious periods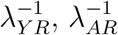 for the symptomatic and asymptomatic sub-populations, weighted with the probabilities of becoming symptomatic (1 − *α*) or asymptomatic *α* upon infection.

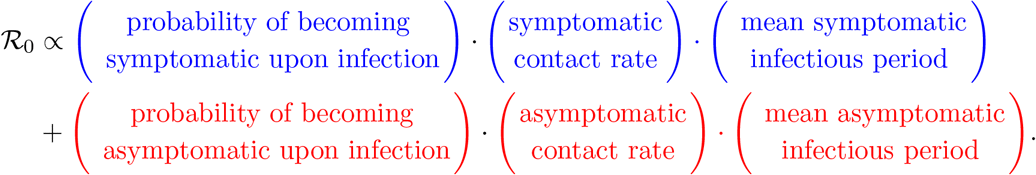

Estimations of the basic reproduction number, 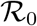, vary on a broad range. The initial estimates of the preliminary outbreak dynamics suggested 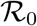 to be in the interval [0.3, 2.38] *(30-38)*. A posterior analysis estimated a median 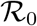 value of 5.7 (95% CI 3.8-8.9) *(39)*; remark 3.1 indicates that the period required to stop the spread of the disease should be at least 58 days with a risk mitigation coefficient of *k* = 0.03.

The reproduction number is not a biological constant corresponding to a given pathogen *(40)*. In reality, the values of 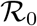 fluctuate with time, and depend on numerous factors. It provides a means to measure the contagiousness of a disease under given circumstances and is utilized by public health authorities to gauge the severity of an outbreak. The design of various public health strategies and measurement of their effectiveness are guided by estimates of 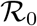. Established outbreaks usually fade provided that interventions maintain 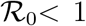. It is defined to be the average number of secondary cases generated by a typical case. A decrease in the susceptible proportion of the population 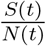 overtime will cause a corresponding decrease in the values of the reproduction number.

The canonical definition of the effective reproduction number 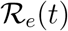 takes into consideration the susceptibility of the population,

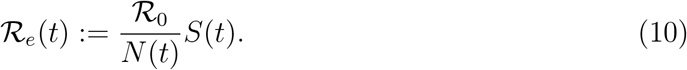

It directly follows by Equation 10 that 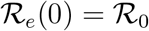, as initially the total human population is assumed to be susceptible. The plot of 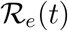 is similar to the plot of the susceptible portion. This is reasonable since Equation 10 implies that 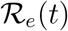 is proportional to *S*(*t*). Since the case fatality ratio *δ* ≈ 0, the total population *N*(*t*) varies little within a tight envelope around the initial susceptible population *S(0)*. This is easily observable upon inspection of the dynamical system given by Equation 4 in Section 2.2, as it is clear that

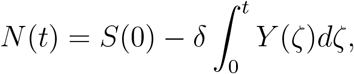

where the function *Y* appearing in the above integral denotes the symptomatic subpopulation.

## 5 Conclusion

As the COVID-19 pandemic evolves, governments around the world are taking drastic steps to limit community spread. This will necessarily dampen the growth of the disease. The parameter *k* introduced in Equation 6 accounts for generalized risk mitigation measures in which there is risk aversion by a substantial portion of the population, thus affecting effective contact rates. We call *k* the *risk mitigation coefficient*. As risk mitigation measures evolve due to government interventions, *k* is adjusted accordingly. In practice, one would set a risk mitigation coefficient corresponding to each mitigation period.

In juxtaposition to the SARS-CoV epidemic of 2003 where only symptomatic individuals were capable of transmitting the disease *(41,42)*, asymptomatic carriers of the SARS-CoV-2 virus may be capable of the same degree of transmission as symptomatic individuals *(11)*. In a public health context, the silent threat posed by the presence of asymptomatic and other undocumented carriers in the population renders the COVID- 19 pandemic far more difficult to control. SARS-CoV-2 is evidently among the more contagious pathogens known, a phenomenon most likely driven by the asymptomatic sub-population.

The calculation of 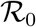 poses significant challenges during the first stages of any outbreak, including the COVID-19 pandemic. This is due to paucity and timing of surveillance data, different methodological approaches to data collection, and different guidelines for testing. Estimates vary greatly: 0.3 *(30)*, 2.28 *(31)*, 2.38 *(8)*, 3.28 *(32)*, and others. The value of 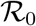 must be understood as a threshold parameter that can be utilized to characterize disease spread. The estimations of 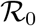 are expected to vary substantially per locality depending on how public health officials communicate the risk to the general public, general beliefs and (dis)information available to the population, and other socioeconomic and environmental factors affecting contact rates.

Different degrees of complication arise due to the presence asymptomatic carriers in an epidemic. If there were relatively few asymptomatic carriers, (for example 1 asymptomatic for every 9 symptomatic), the time to peak could be extended. Many infectious diseases exhibit this behavior and this is far from the worst case scenario; however, controlling the outbreak becomes more challenging. The mid-level situation corresponds to a higher percentage of asymptomatic carries (for example twenty to seventy percent). The relative large number of asymptomatic carriers would accelerate the transmission of the disease, and would make it increasingly difficult to contain. This is the only plausible explanation for the unprecedented speed of propagation of COVID-19. Another possibility is the case where there would be significantly more asymptomatic carriers than symptomatic, (for example 80 times more) *(43)*. Provided that exposure results in immunity, this scenario would be less damaging than the mid-level. If many people get infected, these cases do not develop severity, and exposure confers immunity, this would accelerate the emergence of herd immunity. This could be the most desirable outcome at this point. The scenario most likely occurring is the mid-level; this case creates a protracted epidemic, with a slow build up of herd immunity. This mid-level scenario could result in the need of risk mitigation strategies until there is a vaccine.

The worst-case scenario would consist of the following: (i) mid-level or significantly more asymptomatic carriers than symptomatic, (ii) exposure does not confer immunity, and (iii) a vaccine is elusive as it has been for other coronaviruses that cause the common cold. This unfortunate state of affairs could reduce the life expectancy of our entire species.

## 6 Appendix

**Table 2:**
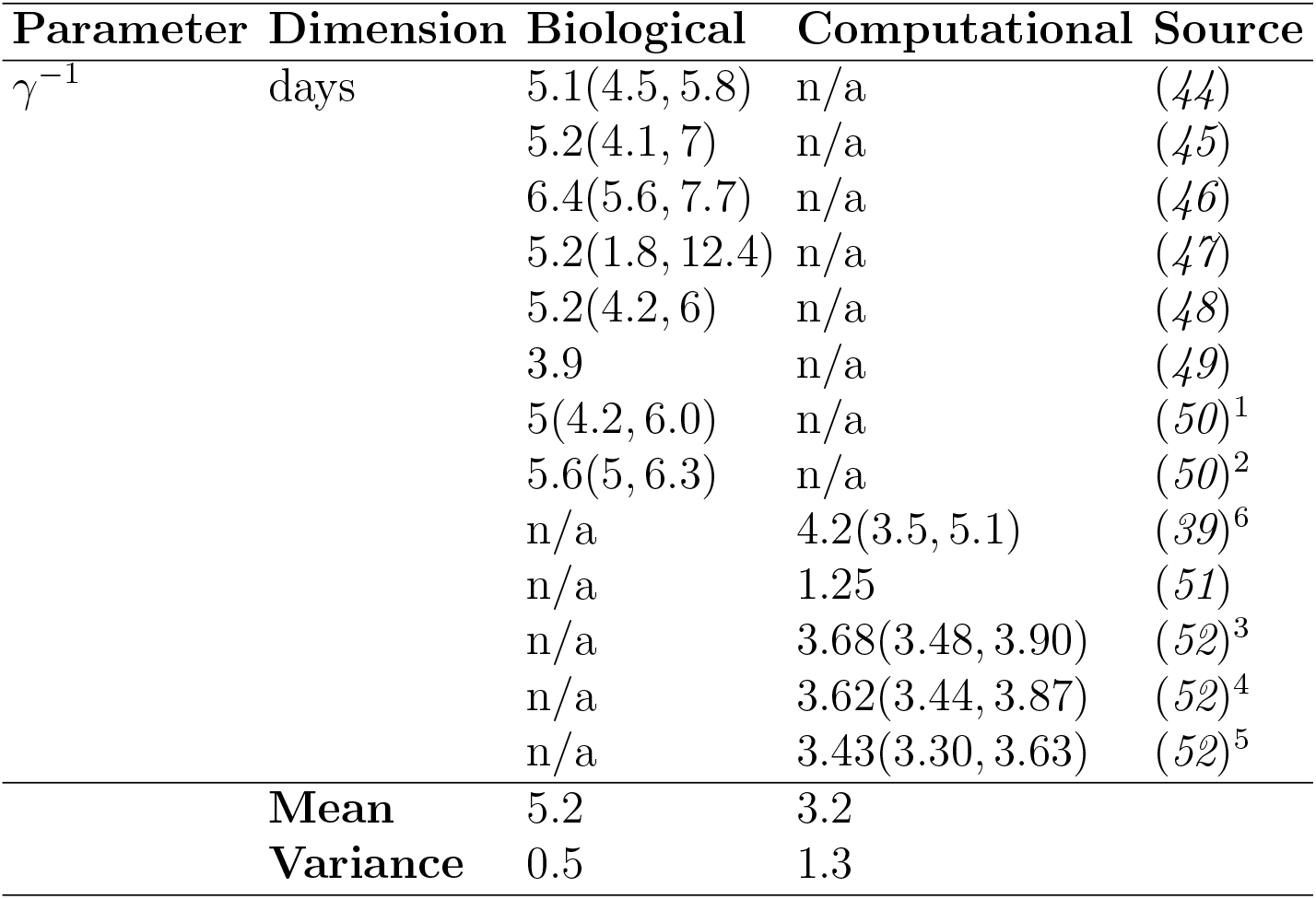
Latent period

**Table 3:** Asymptomatic Proportion

| Parameter | Dimension | Biological | Computational | Source |
| --- | --- | --- | --- | --- |
| $\alpha$ | n/a | 17.9%(15.5%, 20.2%) | n/a | (5) |
|  |  | 33.3% | n/a | (53)<sup>1</sup> |
|  |  | [50%, 75%] | n/a | (7) |
|  |  | 80% | n/a | (54) |
|  |  | n/a | 41.6%(16.7%, 66.7%) | (6)<sup>2</sup> |
|  |  | [80%, 95%] | n/a | (55)<sup>1</sup> |
|  |  | n/a | 86%(82%, 90%) | (52) |
|  |  | 78% | n/a | (56) |
|  | Mean | 60% | 60% |  |
|  | Variance | 10% | 10% |  |

**Table 4:**
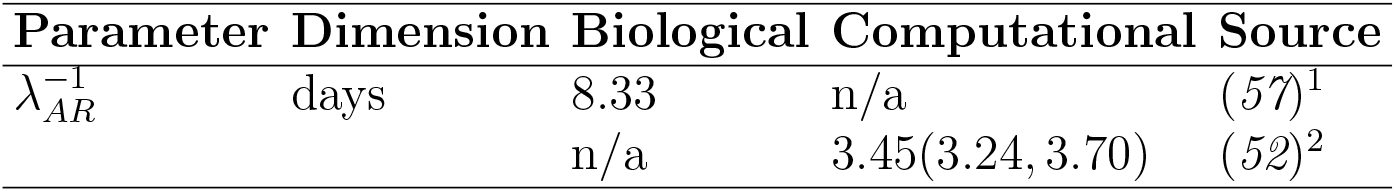
Asymptomatic Infectious Period

**Table 5:**
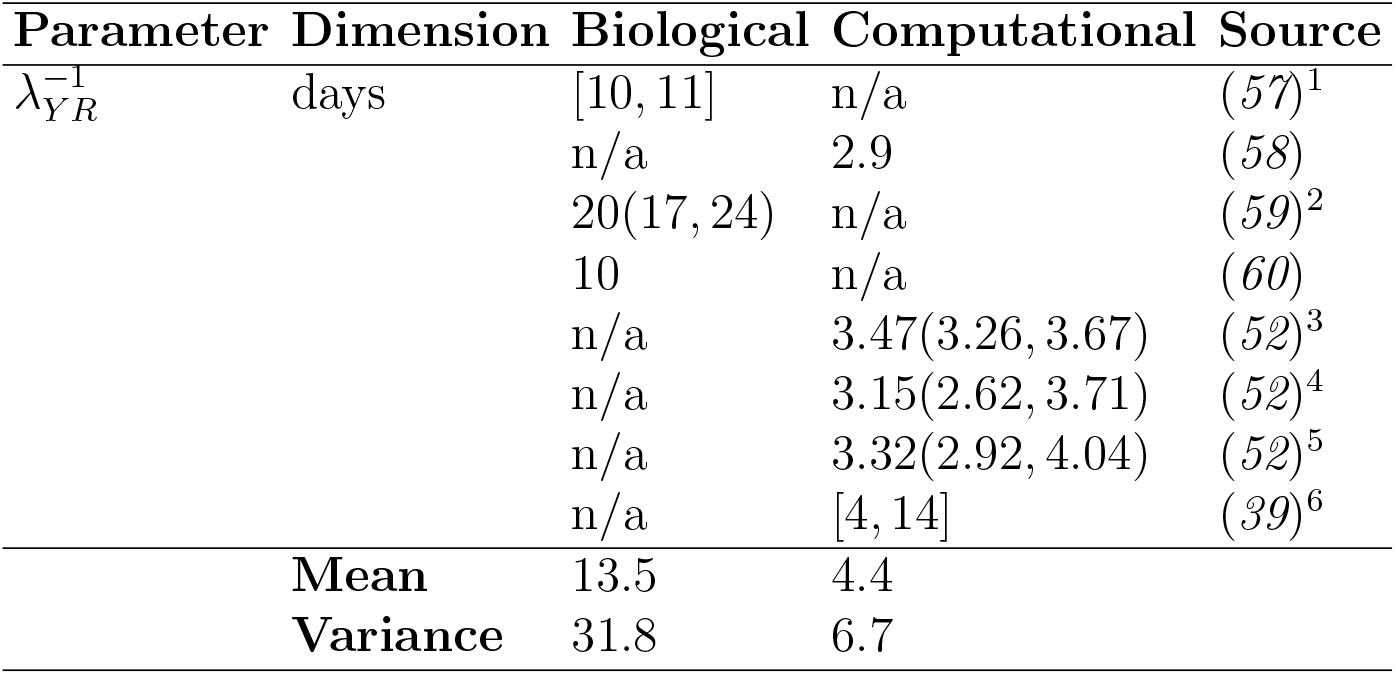
Symptomatic Infectious Period

**Table 6:**
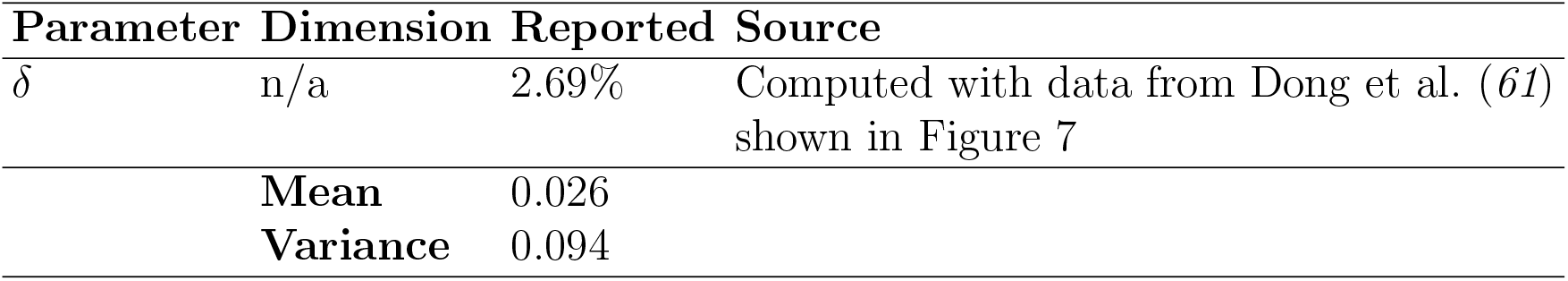
Case Fatality Ratio

**Figure 7:**
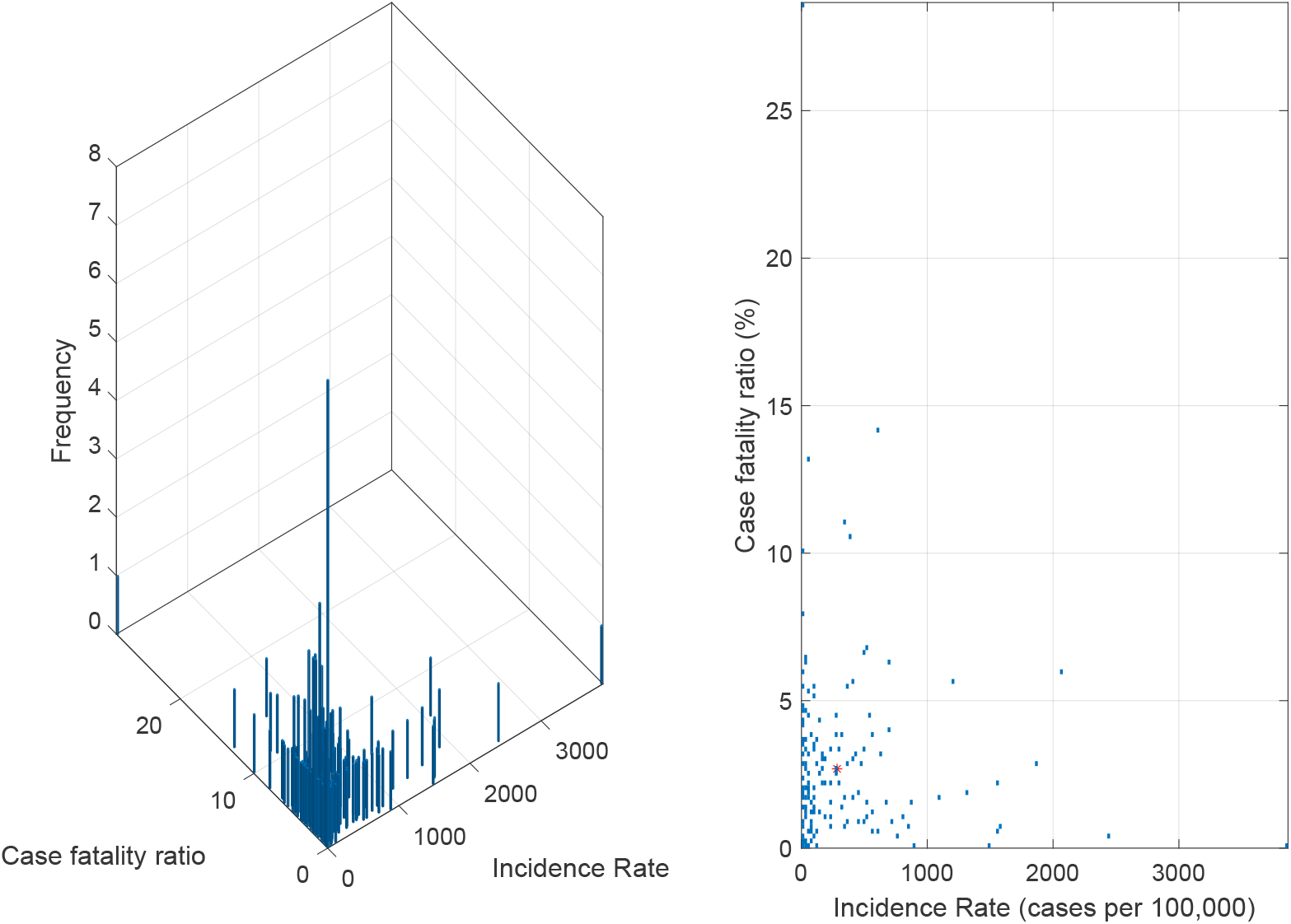
The case fatality ratio *(δ)* is computed as the number of deaths attributed to COVID-19 divided by the number of reported cases on the same period. The mean case fatality ratio from 188 countries as of August 2, 2020, is shown with a red asterisk on the right panel, *δ* = 0.026 (61).

## Data Availability

All data used in this article is available from public sources.

https://mathresearch.utsa.edu/wp/?p=58

## Notes

### Competing Interest Statement

The authors have declared no competing interest.

### Summary of Updates

This is the final version prior to peer-review submission. The immediately previous version contained "lockdown" instead of "risk mitigation" in the title and in the text of the manuscript. We prefer "risk mitigation" because it encompasses a broader range of control strategies, from shelter-in-place to vague guidelines for social distancing.

